# Characterisation of alcohol polygenic risk scores in the context of mental health outcomes: Within-individual and intergenerational analyses in the Avon Longitudinal Study of Parents and Children

**DOI:** 10.1101/2020.07.06.20147231

**Authors:** Kayleigh E Easey, Robyn E Wootton, Hannah M Sallis, Elis Haan, Laura Schellhas, Marcus R Munafò, Nicholas Timpson, Luisa Zuccolo

## Abstract

**Background:** Heavy alcohol consumption often co-occurs with mental health problems; this could be due to confounding, shared biological mechanisms, or causal effects. Polygenic risk scores (PRS) for alcohol use can be used to explore this association at critical life stages.

**Design:** We characterised a PRS reliably associated with patterns of adult alcohol consumption by 1) validating whether it predicts own alcohol use at different life-stages (pregnancy, adolescence) of interest for mental health impact. Additionally, we explored associations of alcohol PRS on mental health phenotypes 2) within-individuals (using own alcohol PRS on own phenotypes) and 3) intergenerationally (using maternal alcohol PRS on offspring phenotypes). We used data from the Avon Longitudinal Study of Parents and Children (ALSPAC) (*n*=960 to 7841). Additional substance abuse behaviours and mental health/behavioural outcomes were investigated (alcohol phenotypes *n*=22; health phenotypes *n*=91). *Findings:* Maternal alcohol PRS was associated with consumption during pregnancy (strongest signal: alcohol frequency at 18 weeks’ gestation: <u>*β*=0.041, 95% CI=1.02 to 1.8)</u>, *p*=1.01×10^−5^, adjusted R^2^=1.16%), offspring alcohol PRS did not predict offspring alcohol consumption. We found evidence for an association of maternal alcohol PRS with own perinatal depression (OR=1.10, 95% CI=0.02 to 0.06, *p*=0.02) and decreased offspring intellectual ability (<u>*β*=−0.209, 95% CI −0.38 to 0.04,</u> *p*=0.016). *Conclusions*: These alcohol PRS are a valid proxy for maternal alcohol use in pregnancy. Offspring alcohol PRS was not associated with drinking in adolescence.

Consistently with results from different study designs, we found evidence that maternal alcohol PRS are associated with both prenatal depression and decreased offspring intellectual ability.

## Introduction

Alcohol use frequently co-occurs with mental health problems (Jané-Llopis & Matytsina, 2006; Mamluk et al., 2020; Skogen et al., 2014), but we do not yet fully understand the nature and extent of these associations. Previous research has implicated two critical life stages. First, during adolescence and early adulthood, when individuals often experience the onset of mental health problems (Kessler et al., 2007) and engage in more risky drinking behaviours (Heron et al., 2012; Marshall, 2014). Second, during pregnancy, as worse mental health outcomes are observed in offspring prenatally exposed to alcohol (Easey, Dyer, Timpson, & Munafò, 2019; Easey, Timpson, & Munafò, 2020; Mamluk et al., 2017). One way to explore the association between alcohol use and mental health outcomes, is using polygenic risk scores (PRS). PRS comprised of genetic variants associated with alcohol use, can act as a proxy for alcohol consumption, and reduce bias from confounding compared with observational evidence (Davey Smith et al., 2007). When exploring the association between alcohol PRS and mental health phenotypes, we can estimate both ‘within-individual’ associations between own alcohol PRS (maternal and then offspring) on own mental health phenotypes (maternal and offspring, retrospectively), as well as investigating ‘intergenerational’ associations between maternal alcohol PRS and offspring mental health phenotypes, to further understand these critical time points.

Genetic variants have been identified that are associated with increased alcohol consumption in adults (Liu et al., 2019), but to date there have been no genome-wide association studies (GWAS) of alcohol consumption in pregnant women or adolescents. We cannot simply assume that the same genetic variants will be associated with alcohol consumption throughout the life course. For example, maternal metabolism is altered during pregnancy to meet the physiological demands on the mother, as well as to promote healthy development of the fetus (Burd, Blair, & Dropps, 2012; Hadden & McLaughlin, 2009; Shankar, Ronis, & Badger, 2007). Furthermore, patterns of drinking during adolescence have been shown to differ compared to adult drinking (Casswell, Pledger, & Pratap, 2002). Therefore, in order to conduct PRS analyses at these critical time points, we must first validate the genetic variants identified in the general population for use in pregnant women and adolescents. For example, the *ADH*_*1*_*B* gene (involved in metabolizing ethanol and linked with alcohol dependence in adults (Bierut et al., 2012) has been shown to also associate with alcohol consumption before and during pregnancy (Zuccolo et al., 2009).

In this paper, our first aim was to validate all genome-wide significant variants associated with alcohol consumption in the most recent GWAS (Liu et al., 2019) for use during pregnancy and adolescence. In the second stage of our analysis, we used these PRS for alcohol consumption to investigate the association between alcohol consumption and mental health. We did this in two ways: first, we explored if maternal and offspring own alcohol PRS were associated with own mental health (within-individual) using a targeted Mendelian randomization phenome-wide association study (MR-PheWAS) design (aim 2). We then examined the intergenerational association between maternal alcohol PRS and offspring alcohol and mental health phenotypes (aim 3). If associations between maternal alcohol PRS and offspring phenotypes are due to shared genetic contributions to both exposure and outcome variables, we might expect weaker associations for maternal alcohol PRS and offspring phenotypes, compared to own alcohol PRS and mental health phenotypes (in either mothers or offspring). This is because offspring share 50% of their variegated genotype with their mother, and we would therefore expect the observed effects to be roughly half the size of those in the mother’s analyses in the presence of a causal intergeneration effect. The same mental health constructs were available throughout the life course, starting from early life (7-10 years), and continuing into adolescence/early adulthood when alcohol use is likely to commence (13-24 years). This allowed us to interpret analyses using outcomes from the early life period as a negative control, since alcohol consumption had not taken place yet and therefore these associations were unlikely to be mediated by child’s own alcohol initiation and use. This study used longitudinal data on multiple generations (mother and offspring) and at different timepoints available from the Avon Longitudinal Study of Parents and Children (ALSPAC). Our analyses were hypotheses generating for potential causal effects, using a targeted MR-PheWAS approach

## Methods

### Study population

The Avon Longitudinal Study of Parents and Children (ALSPAC) is an ongoing population-based study, which recruited pregnant women residing in Avon, UK, with expected dates of delivery between April 1, 1991, and December 31, 1992. The core sample consisted of 14,541 pregnant women, of which 14,062 were live births and alive at 1 year of age.

Participants have been regularly followed up through clinic visits and questionnaires (see supplementary Methods). Detailed information about ALSPAC is available on the study website which includes a fully searchable data dictionary of available data (http://www.bris.ac.uk/alspac/researchers/data-access/data-dictionary). The phases of enrolment are described in more detail in the cohort profile paper and its update (Boyd et al., 2013; Fraser et al., 2013; Northstone et al., 2019).

### Genotyping and quality control

Children from the ALSPAC cohort were genotyped using the Illumina HumanHap550 quad chip genotyping platforms. Mothers from ALSPAC were genotyped using the Illumina human 660W-quad array at Centre National de G*énotypage* (CNG). For further detail on this please see Supplementary Methods, and the methods previously described by Paternoster and colleagues, and Taylor and colleagues (Paternoster et al., 2011; Taylor et al., 2018).

### Alcohol polygenic risk score

A recent large scale GWAS (Liu et al., 2019) used 941,280 individuals and identified 99 SNPs related to the number of alcoholic drinks consumed per week using a threshold of 5×10^−8^, explaining 2.5% of the variance in drinks per week. A total of 34 cohorts were meta-analysed and drinks per week was broadly defined as the average number of alcoholic drinks (aggregated across all alcohol types) consumed per week (see Supplementary Methods). We calculated a PRS for alcohol use based on these 99 conditionally independent genome-wide significant SNPs (Liu et al., 2019) (see Table S1), weights were based on the effect estimates reported in the original publication. The PRS for alcohol use were calculated both for the mothers and offspring using PLINK V1.9. 86 variants and 7921 mothers and 7977 offspring remained after pruning and quality control checks within ALSPAC. The ALSPAC cohort was included within the original GWAS by Liu and colleagues, accounting for 8,913 participants out of a total sample size of 941,280. Previous studies have suggested any bias introduced by this level of overlap would be minimal (Burgess, Davies, & Thompson, 2016). However, as a sensitivity analysis we also ran the main analyses using summary statistics from GSCAN that excluded both ALSPAC and 23andMe.

### Phenotyping

Targeted phenotypes were selected for available alcohol use phenotypes (*n* = 22) and mental health/behavioural phenotypes (*n* = 91) within ALSPAC. For maternal phenotypes (*n =* 30; see Fig. 1), variables were recorded during pregnancy (8 to 32 weeks gestation). For the child’s phenotypes (*n =* 61; see Fig. 2), primary outcome variables were selected as close to age 18 years as possible and negative control variables were selected as close to age 7 years as possible, before alcohol initiation was likely to have occurred. Further details of variable selection are given in Supplementary Methods.

**Fig. 1:**
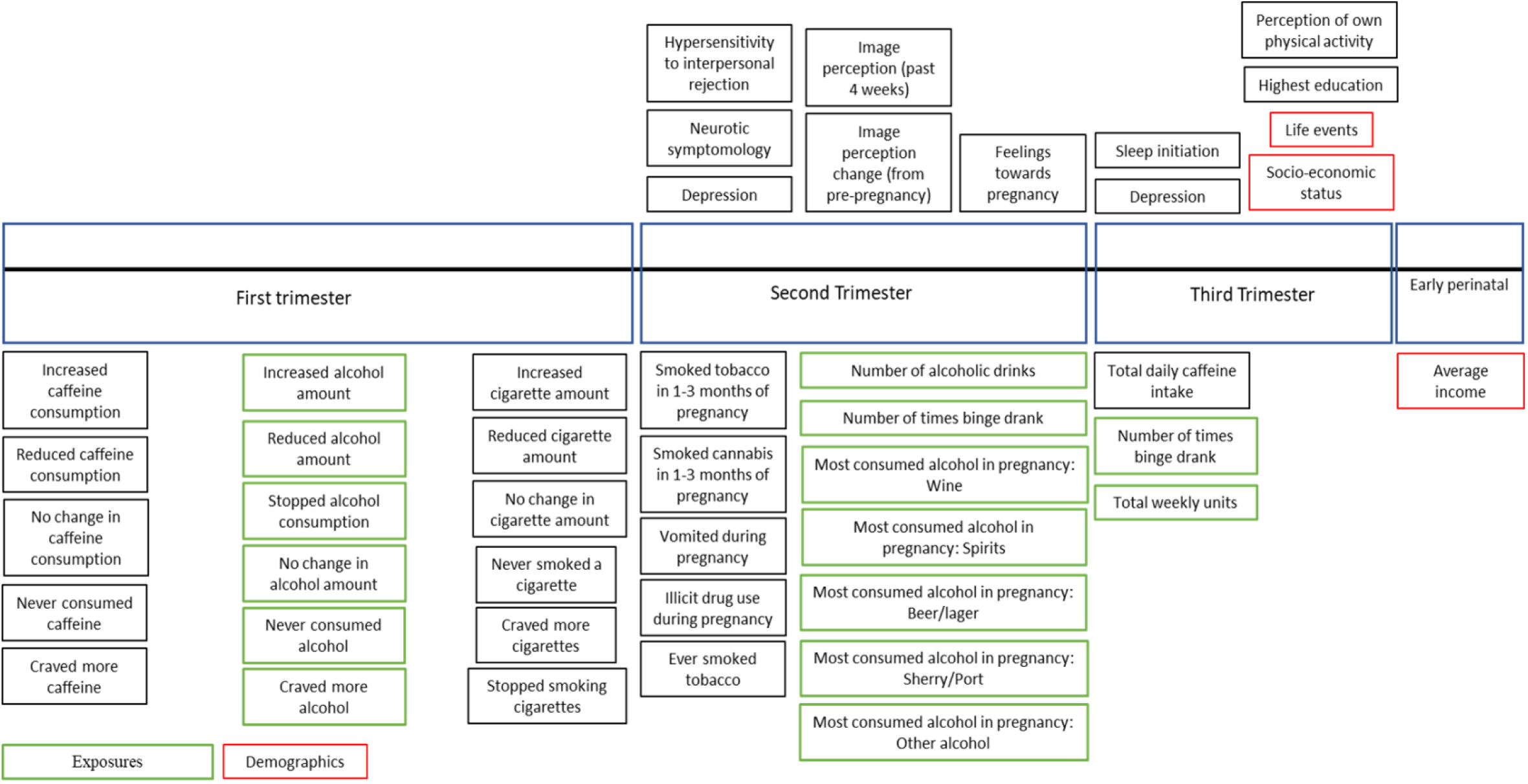
Phenotypes included in targeted MR-PheWAS for mothers during pregnancy

**Fig. 2:**
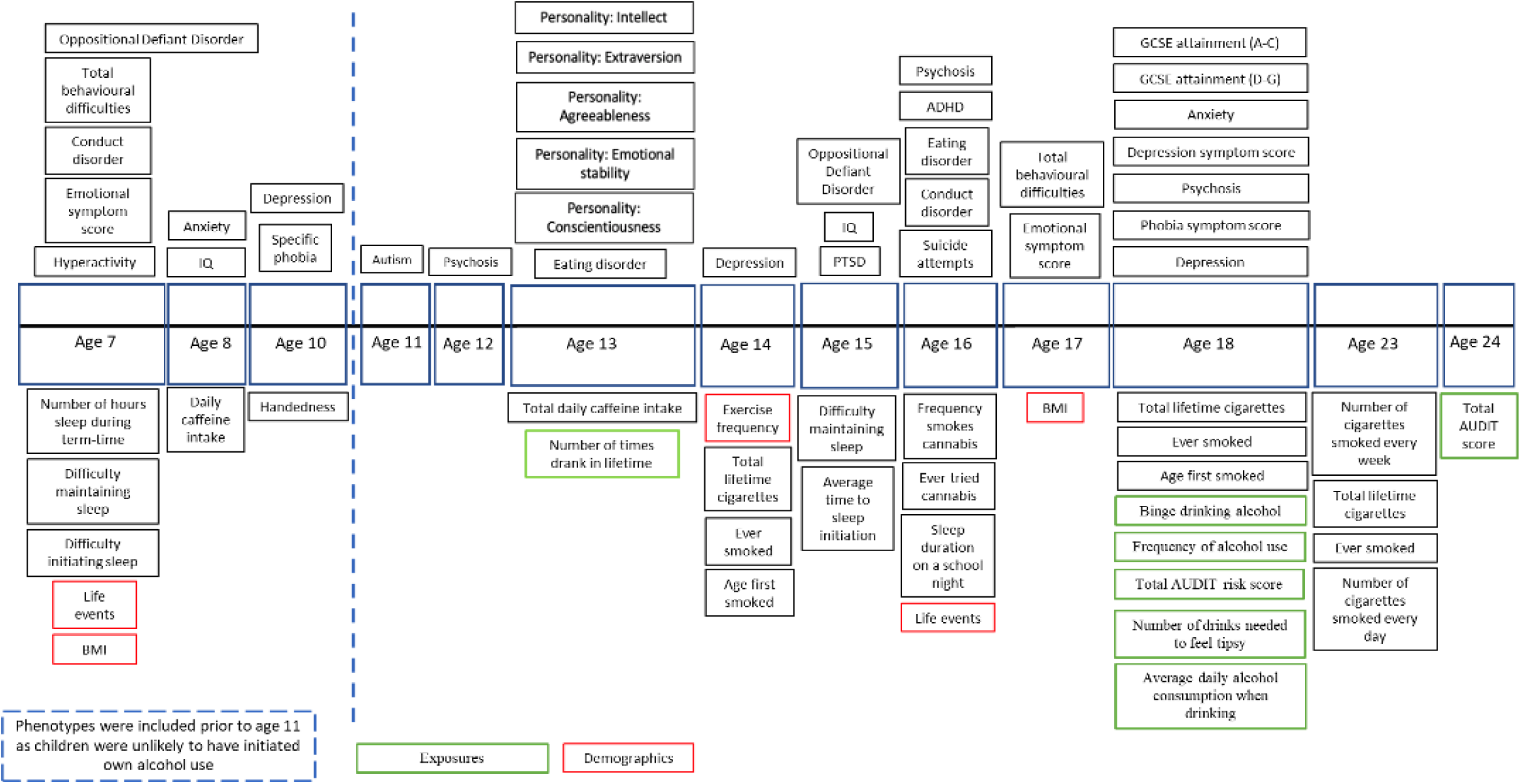
Phenotypes included in targeted MR-PheWAS for children and adolescents

### Statistical analysis

Using Stata version 15.1 (StataCorp, 2017), linear and logistic regression analyses were used to investigate whether own alcohol PRS were associated with 1) alcohol consumption for mothers during pregnancy and in adolescence for offspring, and 2) mental health phenotypes (for mothers during pregnancy, and for children both pre-alcohol use around ages 7-10 years, and post-alcohol use around ages 13-24 years). In addition, similar analyses were used to investigate whether maternal alcohol PRS was associated with 3) offspring alcohol and mental health phenotypes (to test for possible intergenerational effects). All analyses were performed separately in children and mothers. Analyses were adjusted for sex (in the children only), age (at questionnaire completion or clinic attendance), and the first 10 ancestry-informative principal components. We aimed to identify possible causal consequences of alcohol consumption using a targeted MR-PheWAS approach, focusing on genotype-outcome estimates. Mendelian randomization (MR) uses a genetic instrument as a proxy for the exposure of interest (alcohol consumption), to estimate possible causal effects between the exposure and an outcome. We therefore used only genome-wide significant SNPs to construct our PRS, because genetic instruments require specificity over predictive value. In a targeted MR-PheWAS, we expand beyond a single outcome, to over 90 outcomes related to mental health. This approach is hypothesis generating, and any associations identified are suggestive of possible causal effects but should be followed up using formal MR analyses to test for pleiotropy, where a genetic variant influences multiple causal pathways. Pleiotropy is not formally tested within our analyses, apart from the use of a negative control analysis where we used outcomes from early life events occurring before alcohol initiation was likely to have begun, and associations were unlikely to be mediated by child’s own alcohol use. Sensitivity analyses were conducted using Bonferroni-corrected p-values (see Supplementary Methods). For Bonferroni correction, we used an α value of 0.05 / number of tests within each analysis (for categories of phenotypes, see Supplementary Methods). However, Bonferroni correction is a very conservative test, and it was not unexpected that our sample size might not be sufficient to provide statistical evidence of effects when testing multiple highly correlated traits (therefore not independent tests).

Bonferroni correction is more stringent and too conservative in the presence of correlated traits such as the phenotypes used in the current study. Due to the conservative nature of Bonferroni correction when tests are not independent, permutation tests were also conducted accounting for any degree of correlation between the variables in each test.

Three separate Miami plots show associations for 1) validating alcohol PRS at different life-stages (in pregnancy and adolescence), and associations of alcohol PRS and offspring substance use/mental health phenotypes 2) within individuals and 3) intergenerationally (Fig. 3-5). These figures indicate the direction of association by multiplying the −log_10_ *p* by −1 for results of a negative β: sgn(β)(−log_10_ *p*).

**Fig. 3:**
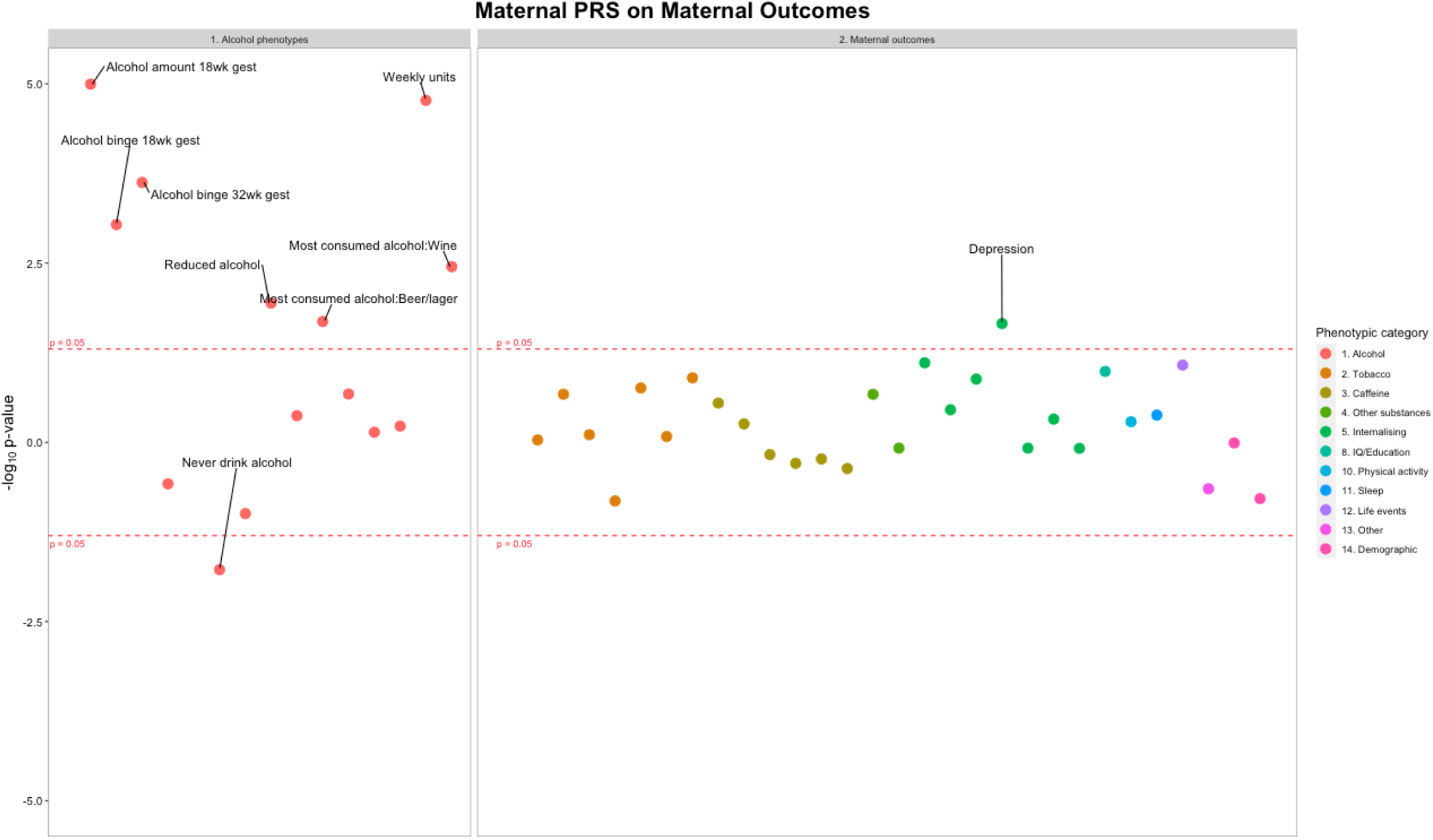
Associations between maternal alcohol PRS and maternal alcohol exposures and mental health phenotypes. Each datapoint represents an individual phenotype, colour coded by phenotypic category. Y axis: sgn(β)(−log_10_ *p*). See Tables S3 and S4 for sample sizes.

Follow up analyses were conducted to investigate associations of maternal alcohol PRS with of maternal lifetime drinking (i.e., outside of pregnancy), including maternal alcohol consumption frequency when offspring were aged 4 and 8 years, number of occasions mothers binge drank, and maternal AUDIT risk.

As a further sensitivity analysis, we recreated the PRS using summary statistics excluding both ALSPAC and 23andMe and repeated our main analyses on this PRS. This did not substantially change our findings, see Supplementary Tables 10-12.

## Results

Sample sizes ranged between 2,540 and 7269 for analyses of maternal alcohol PRS and maternal phenotypes, between 1,035 and 7,841 for offspring alcohol PRS and offspring phenotypes, and between 960 and 7,727 for intergenerational analyses of maternal alcohol PRS and offspring phenotypes, see Supplementary Tables for sample sizes for individual analyses.

Of the mothers who responded to the questionnaire concerning alcohol use at 18 weeks gestation, 2% reported never consuming alcohol during this pregnancy, while 98% reported that they had consumed alcohol and 18% reported binge drinking within pregnancy. *Aim 1) Alcohol use PRS and alcohol phenotypes*. Maternal alcohol PRS previously identified in the general population were shown to be associated with maternal alcohol use during pregnancy (see Fig. 3). Each SD increase in genetic score for alcohol consumption was associated with a 0.04 unit increase in the number of drinks consumed each week at 18 weeks gestation (95% CI 0.02 to 0.06, *p* = 1.01 × 10^−5^, *n =* 7,185), 0.03 unit increase in the number of days mothers binge drank at 18 weeks gestation (95% CI 0.01 to 0.05, *p* = 9.19 × 10^−4^, *n* = 7,171), 0.04 unit increase in the number of days mothers binge drank at 32 weeks gestation (95% CI 0.02 to 0.06, *p* = 2.37 × 10^−4^, *n* = 5,324) and 0.25 unit increase in the in the number of drinks consumed each week at 32 weeks gestation (95% CI 0.04 to 0.36, *p* = 1.70 × 10^−5^, *n =* 4,294) (Fig. 3). There was little evidence that offspring PRS for alcohol use was associated with any of the offspring alcohol measures at ages 13 or 18 years (Fig. 4).

**Fig. 4:**
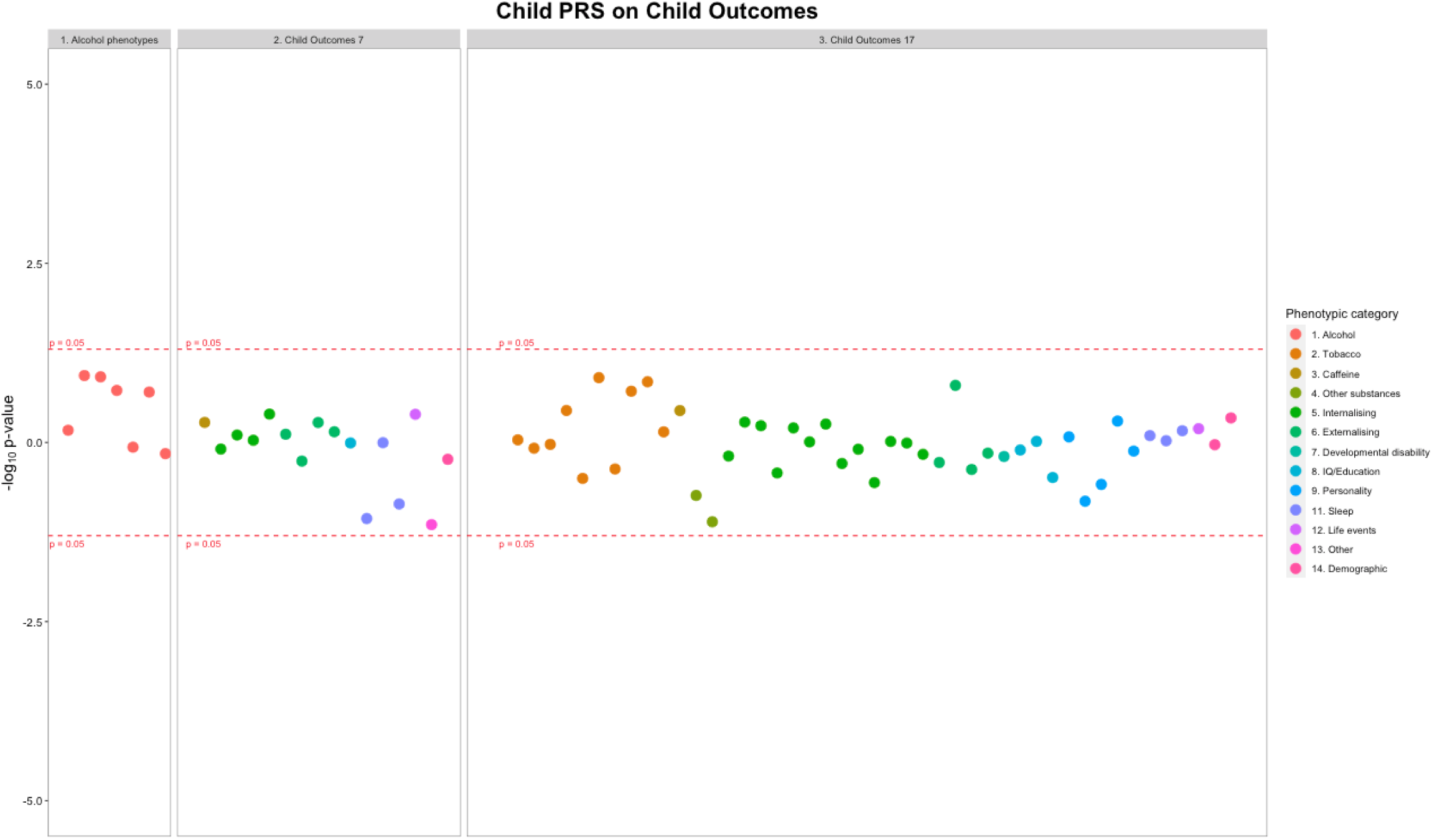
Associations between offspring alcohol PRS and offspring alcohol exposures, mental health phenotypes at pre-drinking age and drinking initiation age. Each datapoint represents an individual phenotype, colour coded by phenotypic category. Y axis: sgn(β)(−log_10_ *p*). See Tables S3 and S5 for sample sizes.

*Aim 2) Alcohol use PRS and mental health phenotypes (during pregnancy, age ~7 and age ~18)*. Maternal alcohol PRS was associated with increased maternal depression at 32 weeks gestation (OR = 1.09, 95% CI = 1.02 to 1.18, *p* = 0.022, *n* = 6,751), see Table S4. There was no clear evidence of association between maternal alcohol PRS and any of the other mental health phenotypes (Fig. 3). There was little evidence that offspring alcohol PRS were associated with any of the child mental health phenotypes (see Table S5).

*Aim 3) Intergenerational. Maternal alcohol PRS and offspring alcohol and mental health phenotypes (age ~7 and age ~ 18)*. Maternal alcohol PRS were associated with increased offspring AUDIT total score (β = 0.184, 95% CI = 0.02, to 0.35, *p* = 0.028, *n* = 2,516) (Table S3, and Fig. 5). Of the measured offspring phenotypes, maternal alcohol PRS were associated with decreased scores in intellectual ability at age 13 (β = −0.209, 95% CI = −0.38 to −0.04, *p* = 0.016, *n* = 3,956, see Tables S3 and S6). Maternal alcohol PRS showed little evidence of association with any of the other offspring outcomes (see Table S6).

**Fig. 5:**
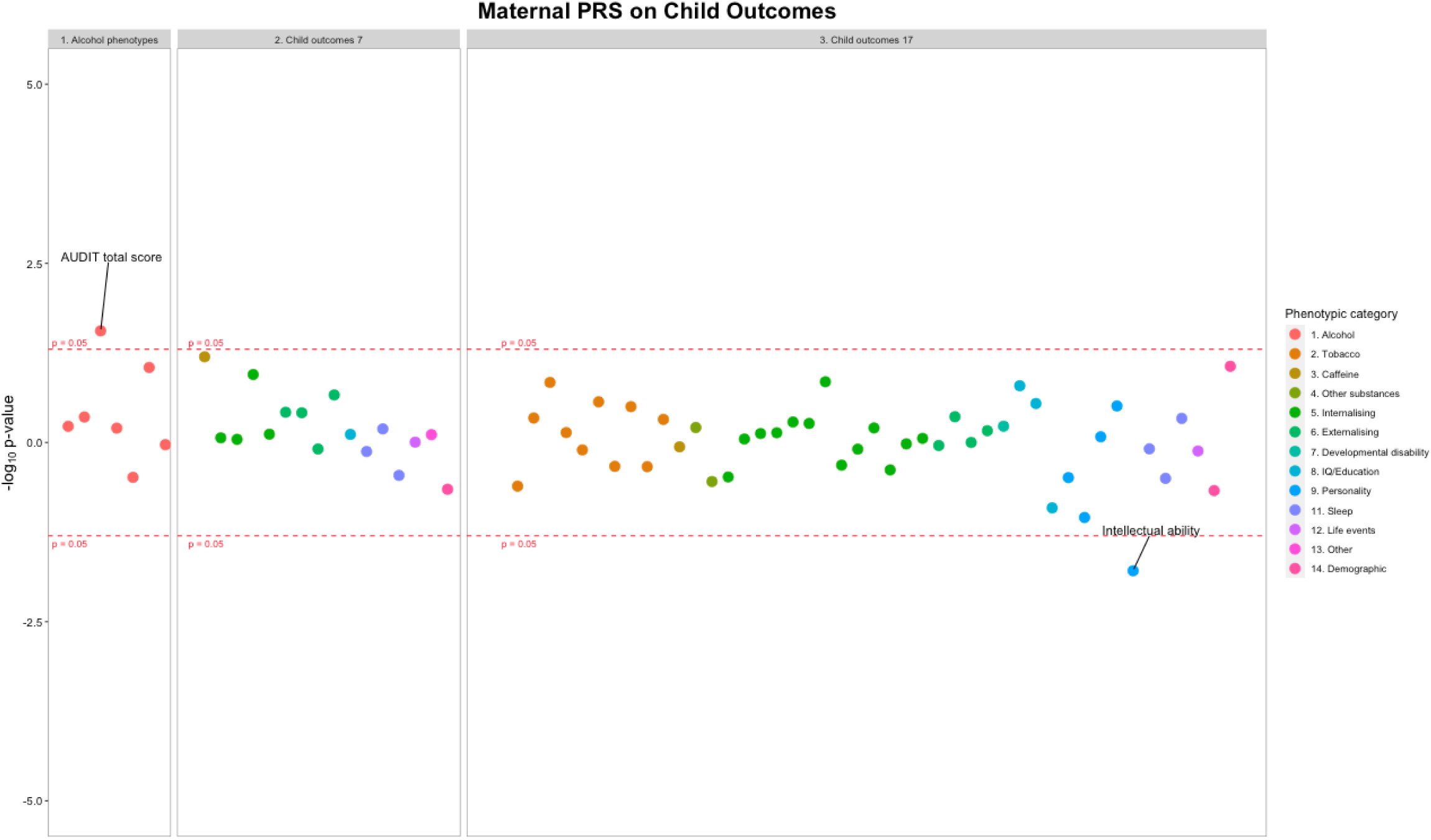
Associations between maternal alcohol PRS and offspring alcohol exposures and mental health phenotypes at pre-drinking age and drinking initiation age (intergenerational). Each datapoint represents an individual phenotype, colour coded by phenotypic category. Y axis: sgn(β)(−log_10_ *p*). See Tables S3 and S6 for sample sizes.

Follow up analyses of measures of maternal drinking outside of pregnancy showed each SD increase in maternal alcohol PRS was associated with a 0.03 unit increase in the number of alcoholic units consumed daily when offspring were age 4 (95% CI 0.01 to 0.05, p = 8.0 × 10^−4^, *n* = 2,707), and 0.05 at age 8 (95% CI 0.02 to 0.08, p = 8.9 × 10^−4^, *n* = 2,707), as well as a 0.07 unit increase in the number of days mothers binge drank when offspring were age 5 (95% CI 0.04 to 0.10 p = 5.5 × 10^−6^, *n* = 4,866). Maternal alcohol PRS was also associated with increased maternal AUDIT total score when offspring were age 18 (β = 0.03, 95% CI = 0.01 to 0.05, p = 0.006).

*Sensitivity analyses*. After permutation analyses, the strength of evidence persisted for maternal alcohol PRS associating with maternal depression at 32 weeks gestation: *p =* 0.016. Whereas the intergenerational association between maternal PRS and offspring phenotypes was attenuated (see Tables S7 to S9).

## Discussion

We have shown that PRS for alcohol consumption reliably associates with consumption in pregnancy, but not in adolescence/young adulthood. This suggests that alcohol PRS derived in the general population is associated with consumption patterns during pregnancy and can be applied in future epidemiological analysis focused on this timing for exposure. Although we couldn’t find conclusive statistical evidence for an effect of the alcohol PRS in predicting consumption in adolescence, the possibility of a false negative remains, due to limited statistical power. Other reasons for this finding include a true difference in genetic susceptibility for earlier compared with later drinking behaviour. The discovery sample for the SNPs included in our PRS was a collection of (mostly) middle and older age individuals (Liu et al., 2019), with average age much older compared to the adolescents of our target sample (ALSPAC). For example, in adolescents and younger adults, drinking patterns are not yet fully established and may be more subject to social influences (Maggs & Schulenberg, 2004). Previous studies have shown that alcohol use often increases over time from adolescence to adulthood (Casswell et al., 2002; Paavola, Vartiainen, & Haukkala, 2004). We also found that maternal alcohol PRS was associated with maternal alcohol use phenotypes outside of pregnancy, replicating recent results from heterogeneous populations, and explaining up to 3.5% of variance in alcohol use (Barr et al., 2019; Chang et al., 2019) (Barr et al., 2019; Chang et al., 2019). However, much of the previous research using the drinks per week PRS (Liu et al., 2019) in a general sample of adults, has focused on alcohol use disorders as opposed to general (non-problematic) drinking. The current study adds to this literature by further validating the SNPs from Liu and colleagues as expected in a general adult sample. We subsequently investigated associations between both maternal and offspring PRS for increased alcohol consumption, and multiple mental health outcomes. In within-individual analyses we found evidence of association between maternal PRS and increased maternal depression at 32 weeks gestation. Alcohol abuse is shown to have high comorbidity with mental health problems (Burns & Teesson, 2002; Kessler et al., 1997), and these results may suggest a potential causal effect of alcohol consumption on maternal depression, or a shared biological contribution to both alcohol consumption and mental health. MR studies have previously identified a causal role of genetic liability for major depression and alcohol dependence (Polimanti et al., 2019; Zhou et al., 2017). Our finding is in line with this literature, and it extends it to lower non-pathological alcohol use levels in a general population of pregnant women.

The offspring alcohol PRS was also not shown to be associated with any of the mental health outcomes in the offspring subpopulation, at ages ~7 and ~18 years. The outcomes at age 7 provided a negative control, because the offspring are unlikely to have started consuming alcohol themselves. Therefore, any associations are likely to be due to horizontal pleiotropy or maternal drinking, rather than the offspring’s own alcohol use. However, as the offspring alcohol PRS did not predict offspring alcohol consumption, we cannot make inferences about the association between alcohol use and mental health. The sample sizes for offspring phenotypes were also relatively small as they were mostly collected around age 18, when the cohort has experienced significant attrition. Larger sample sizes would increase statistical power and potentially identify weaker associations.

We also investigated the effect of maternal alcohol PRS on offspring phenotypes to elucidate potential intergenerational associations. Within the intergenerational analyses, maternal alcohol PRS were associated with offspring increased risk of hazardous alcohol use (AUDIT), but no other alcohol measures. The association with hazardous use could be due to genetic nurture, with offspring being more exposed to a parent engaging in high levels of alcohol use. The lack of association with other outcomes could again be due to the younger age of the adolescents having not yet fully established their drinking behaviour, or the smaller sample size and consequently reduced statistical power. Maternal alcohol PRS was also associated with decreased scores for intellectual ability. This could be either due to a causal intrauterine effect, offspring’s own alcohol use (which is unlikely as we did not find an association with offspring alcohol use), or pleiotropic effects. As we had already shown maternal PRS to be a measure of increased maternal alcohol use, this would suggest that offspring of mothers with increased levels of alcohol PRS would have likely been exposed to greater amounts of prenatal alcohol exposure (PAE). Permutation testing attenuated the strength of evidence in support of the intergenerational associations for offspring cognitive outcomes.

We chose to use PRS comprising only genome-wide significant SNPs, because we valued specificity over predictive value for our current research question. We aimed to identify possible causal consequences of alcohol consumption using a targeted MR-PheWAS that could be followed up in downstream MR analyses. Therefore, selecting genome-wide significant SNPs reduced the likelihood of pleiotropic associations and ensured the PRS was more specific to alcohol consumption. We acknowledge that this strategy does not maximise predictive value of the PRS, however, it does improve specificity (for the alcohol phenotype) as well as minimising pleiotropy. This decision was guided by the downstream application of the PRS in the context of causal inference – in particular its employment in MR analyses. Future investigation to formally investigate and detect pleiotropy should be investigated in future studies using 2-sample MR.Key strengths of this study are the validation of the alcohol PRS in pregnant women, and its novel, proof-of-principle application to intergenerational analyses. However, several limitations should also be considered when interpreting these results. First, the small sample sizes reduced our power to detect true associations. As PRS typically only explain a small proportion of phenotypic variance, even a moderately large sample as the ALSPAC study is not sufficient on its own to rule out moderate or small effect sizes in this context. Our results therefore warrant replication. Future follow-up studies should therefore use cohorts with larger sample sizes to address how alcohol PRS may influence mental health trajectories within the two generations of pregnant mothers and their offspring. Second, we did not conduct longitudinal analyses. Third, there was modest sample overlap between ALSPAC and the GWAS in which the SNPs for alcohol consumption were identified (13); the GWAS included 8913 participants from ALSPAC out of a total sample size of 941,280. However, this was unlikely to introduce substantial bias as the size of this overlap compared to the total included in the GWAS is minimal. As suggested by Burgess and colleagues (Burgess et al., 2016), we ran a sensitivity analysis to assess bias due to sample overlap. This showed no evidence of bias, with the estimates for all analyses largely unchanged. Previous studies have used similar sensitivity analyses when faced with sample overlap, and typically also find no evidence of bias when sample overlap is modest (Linden et al., 2020).

The current methods and data do not allow us to draw firm conclusions regarding the mechanisms underpinning the observed associations, and this was never the study’s intention. Taken together with prior evidence from different study designs, which are robust to different biases, our intergenerational results point to a causal effect of mothers’ alcohol PRS on offspring outcomes. In particular, this is consistent with a body of literature demonstrating adverse effects of PAE on offspring cognition (Mamluk et al., 2020). However, increased post-natal maternal alcohol use may also influence offspring outcomes, for example via how the mother interacts with or parents her child or environmental pathways (Lieb et al., 2000). In contrast, the likelihood that these findings could be due to horizontal pleiotropy, or shared genetic liability between increased alcohol use and cognition is low. This is because on the one hand, we did not observe any associations between child’s own alcohol PRS and child cognition, and on the other, the offspring outcomes where there was evidence of an association with maternal PRS occurred before the offspring were likely to have begun consuming alcohol.

Our findings validate the alcohol PRS as a reliable predictor for maternal alcohol consumption during pregnancy, but caution that the same PRS might not be a suitable proxy for consumption in adolescence. The additional analyses we carried out as part of our within-individual and intergenerational analyses (aims 2 and 3) replicated well-known epidemiological associations (e.g., of maternal alcohol use in pregnancy and lower offspring cognition), and extended the known association of alcohol abuse and depression (Grant & Harford, 1995) to lower levels of drinking, and within pregnancy. Characterising alcohol PRS in the context of alcohol and mental health outcomes in this richly characterised multigenerational cohort should therefore be viewed as more than simply a proof-of-principle approach. Nevertheless, our results should be interpreted in the context of the study’s limitations, and in particular that this approach alone cannot fully disentangle if associations are evidencing causal pathways or are due to pleiotropy. Follow-up analyses should be conducted within larger cohorts and focused on causal inference, for example, detailed MR analyses.

## Data Availability

Data is available through application to the research executive of ALPSAC. Please note that the study website contains details of all the data that is available through a fully searchable data dictionary and variable search tool: http://www.bristol.ac.uk/alspac/researchers/our-data/.

## Acknowledgments

We are extremely grateful to all the families who took part in this study, the midwives for their help in recruiting them, and the whole ALSPAC team, which includes interviewers, computer and laboratory technicians, clerical workers, research scientists, volunteers, managers, receptionists and nurses. This work was supported by the UK Medical Research Council and the Wellcome Trust (Grant ref: 102215/2/13/2) and the University of Bristol provide which provide core support for ALSPAC. This work was performed in the UK Medical Research Council Integrative Epidemiology Unit, jointly funded by the University of Bristol and the UK MRC, in which MRM leads one of the programmes (MC_UU_00011/7). The MRC also funded KEE’s PhD studentship. LZ was supported by a UK Medical Research Council fellowship (grant number G0902144). This research was also supported by the NIHR Bristol Biomedical Research Centre at University Hospitals Bristol NHS Foundation Trust and the University of Bristol. The views expressed in this publication are those of the authors and not necessarily those of the NHS, the National Institute for Health Research or the Department of Health and Social Care. NJT is a Wellcome Trust Investigator (202802/Z/16/Z), is the PI of the Avon Longitudinal Study of Parents and Children (MRC & WT 102215/2/13/2), is supported by the University of Bristol NIHR Biomedical Research Centre (BRC-1215-20011), the MRC Integrative Epidemiology Unit and works within the CRUK Integrative Cancer Epidemiology Programme (C18281/A19169). LS and EH are funded by CAPICE (Childhood and Adolescence Psychopathology: unravelling the complex etiology by a large Interdisciplinary Collaboration in Europe) project, funded by the European Union’s Horizon 2020 research and innovation programme, Marie Sklodowska Curie Actions – MSCA-ITN-2016 – Innovative Training Networks under grant agreement number 721567. The UK Medical Research Council also funded the most recent collection of alcohol information in the ALSPAC participants (grant number MR/L033306/1. A comprehensive list of grants funding is available on the ALSPAC website (http://www.bristol.ac.uk/alspac/external/documents/grant-acknowledgements.pdf). GWAS data was generated by Sample Logistics and Genotyping Facilities at Wellcome Sanger Institute and LabCorp (Laboratory Corporation of America) using support from 23andMe. This publication is the work of the authors and they will serve as guarantors for the contents of this paper.

